# SNP-Associated Differential Methylation in *ARHGEF38:* Insights into Genetic-Epigenetic Interactions

**DOI:** 10.1101/2025.02.28.25322876

**Authors:** Emese H.C. Kovács, Lucas G. Casten, Niamh Mullins, Jenny Gringer Richards, Aislinn J. Williams, John A. Wemmie, Vincent A. Magnotta, Jess G. Fiedorowicz, Jacob Michaelson, Marie E. Gaine

## Abstract

**Objective:** Associations have been seen between suicidal behavior and differential DNA methylation of certain genes, with one study showing significant hypomethylation of *ARHGEF38* in postmortem brain samples from individuals with bipolar disorder who died by suicide. Our objective was to explore *ARHGEF38* methylation in individuals with bipolar disorder and a history of suicide attempt.

**Method:** With pyrosequencing, we looked at the previously identified region of interest in *ARHGEF38.* We investigated the methylation levels of 3 CpG sites in 47 individuals with bipolar disorder and a history of suicide attempt, 47 individuals with bipolar disorder without a history of suicide attempt, and 47 non-bipolar disorder controls.

**Results:** None of the CpG sites measured had an association between groups, although there were distinct clusters of differential methylation in each group. Applying genotypes of SNPs found in the region of interest, rs2121558 and rs1447093, these clusters showed stepwise methylation at each CpG site, regardless of phenotype.

**Conclusions:** In this relatively small sample size study, differential methylation in *ARHGEF38* was not associated with history of suicide attempt, failing to replicate findings from a related outcome, suicide death. However, we did provide evidence of SNP and DNA methylation interplay in this region. This highlights the potential relevance of considering genetics when interrogating epigenetic mechanisms.

**Highlights:**

- *ARHGEF38* methylation is not associated with bipolar disorder and suicide attempt
- Methylation of *ARHGEF38* is heavily influenced by the presence of SNPs
- Suicide phenotype, genetics, and sample type impact DNA methylation

## 1. Introduction

Bipolar Disorder (BD) is a mood disorder characterized by depressive and (hypo)manic episodes^1^. BD is associated with a high incidence of suicide attempts (SA); attempt estimates range from 10 to 30 times higher than those of the general population^2–5^, and approximately 7.8% of people with BD die by suicide^6^. Suicidal behavior (SB), encompassing both SA and suicide death (SD)^7,8^ may manifest due to biological and environmental factors, as well as personality traits and early-life adversity^9–13^.

DNA methylation (DNAm) has been associated with increased risk for SB. Postmortem brain studies have identified differentially methylated sites in suicide decedents, indicating that this phenotype may be influenced by DNAm^14–18^. This includes Gaine et al., which reported a differentially methylated region within the Rho Guanine Nucleotide Exchange Factor 38 gene (*ARHGEF38)* in the prefrontal cortex of individuals with BD who died by suicide compared to nonpsychiatric controls who died from other causes^18^. This study showed significant 23.4% hypomethylation (FWER=0.009) in SD with BD across four *ARHGEF38* CpG sites. In this follow-up study, we used a cohort of BD subjects with and without SA to investigate if *ARHGEF38* is also differentially methylated in SA.

## 2. Materials and Methods

### 2.1 Sample Collection

Individuals were recruited through the Iowa Neuroscience Institute Bipolar Disorder Research Program of Excellence (BD-RPOE) with inclusion and exclusion criteria published elsewhere^19–25^. History of SA was recorded using the Columbia Suicide Severity Rating Scale (C-SSRS)^26^. Attempts did not have to result in injury or harm but had the potential for injury or harm. C-SSRS responses were used to create two cohorts: BD-I subjects with a history of SA (BDS) and without a history of SA (BDNS). Control participants (CON) did not have BD (or immediate family members with BD) or a history of SA (N=47 for each group). This study was approved by the University of Iowa (IRB #201708703) and all participants provided informed written consent prior to entering the study.

### 2.2 Pyrosequencing

Five hundred nanograms of genomic DNA per sample were bisulfite converted using the EpiTect Fast 96 DNA Bisulfite Kit (Qiagen) or the EZ DNA Methylation Kit (Zymo Research). Bisulfite-converted DNA was amplified with the PyroMark PCR Kit (Qiagen), and two of the three optimized primers designed by Gaine et al.^18^. The third primer could not be consistently optimized. Upon further investigation, a SNP within this CpG site was discovered, rs1447093 (minor allele frequency=0.195), that changes the guanidine of the CpG site to a cytosine, which would explain these inconsistencies. Pyrosequencing was performed on the PyroMark Q48 Autoprep using the Q48 Advanced CpG Reagent Kit (Qiagen).

Methylation percentages that failed quality control for pyrosequencing were imputed using the R package *missRanger*^27^.Quality control for pyrosequencing failed at all three CpG sites, for four samples in the BDS group, five samples in the BDNS group, and three samples in the CON group. This resulted in sample sizes of 43 BDS, 42 BDNS, and 44 CON.

### 2.3 Genotyping

We used the UCSC Genome Browser^28^ to find common SNPs in the area of interest, resulting in three SNPs: rs2121558, rs10516521, and rs1447093. **Figure 1** describes the CpG sites and SNPs investigated in this study.

**Figure 1.**
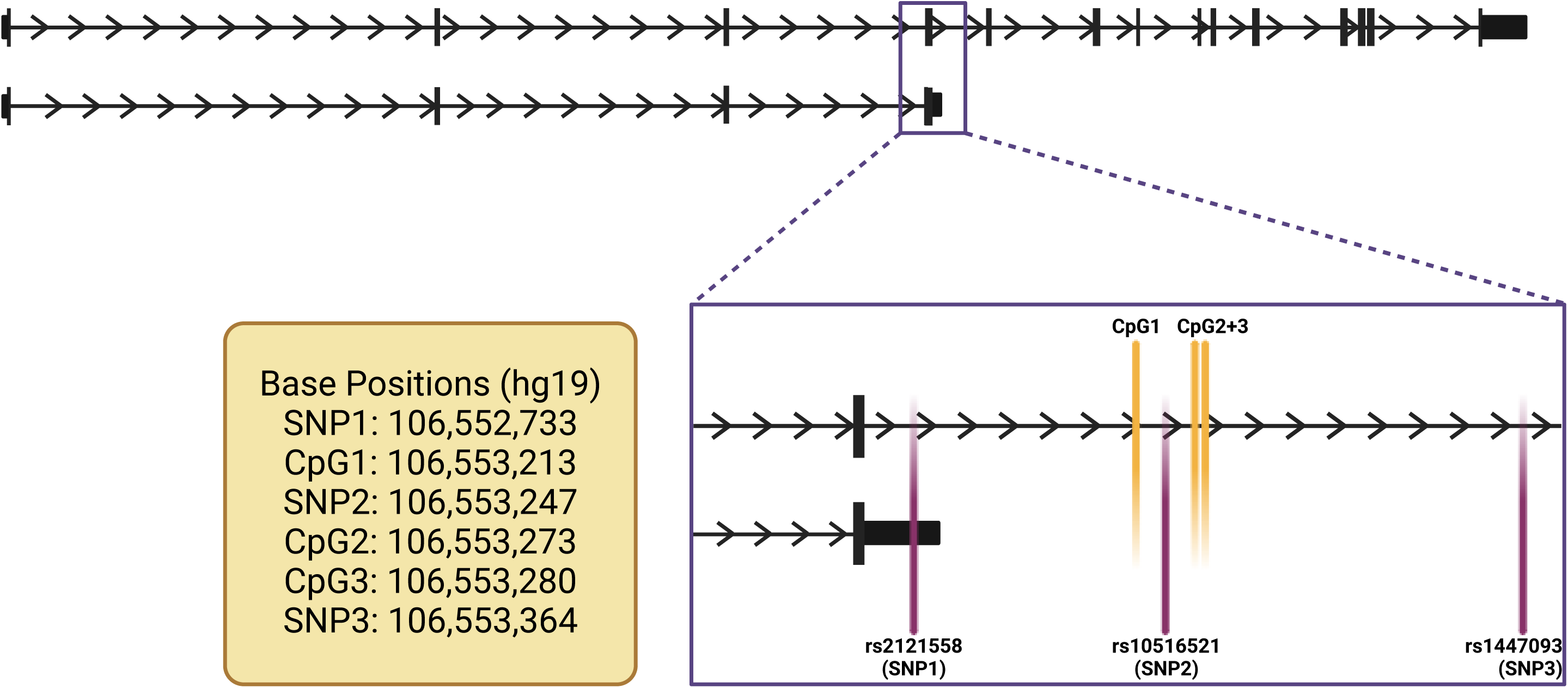
Schematic of *ARHGEF38* and location of SNPs and CpGs analyzed in this study. A representative image not to scale. All SNPs and CpG sites are located on chromosome 4. Figure made in BioRender.

Genetic data generated from PsychArrays was available for some participants from a prior study^29–31^. This work captured 34 of the 44 BDS, 37 of the 44 BDNS, and 36 of the 45 CON participants. Briefly, genotypes were called using Illumina GenomeStudio2.0 with the manufacturer’s provided cluster files. Data was filtered down to SNPs, removing indels and non-biallelic sites using PLINK^32^. The three chosen SNPs were extracted for the appropriate samples.

For samples not analyzed in the PsychArray, we identified the genotype of the samples at these SNPs with TaqMan SNP Genotyping Assays (C 3257733_20, C 30117127_20, C 7627697_10; ThermoFisher). The wet DNA delivery method was used per manufacturer’s instructions, then PCR was performed on the QuantStudio 3 RT-PCR System (ThermoFisher). Eight samples that were previously run on the PsychArray were also run on the TaqMan assays to check assay reliability.

### 2.4 Summary data-based Mendelian Randomization (SMR)

SNPs rs2121558, rs10516521, and rs1447093 were each investigated for putative causal relationships with SA via DNAm using SMR v1.03^33,34^ by integrating methylation quantitative trait loci (meQTLs) from the Brain-mMeta study v2^35^, comprising fetal brain and adult cortex samples measured on the Illumina HumanMethylation450K array, and a European-ancestry SA GWAS^36^. Each SNP was defined as the target SNP for DNAm probes within a ±2Mb window for which the SNP had *P*_meQTL_<5E-08, totaling 9 SNP-probe tests. Statistical significance thresholds were *P*_SMR_<0.005 and *P*_HEIDI_≥0.01 (heterogeneity in dependent instruments test) to ensure SMR assumptions were not violated by horizontal pleiotropy^34^.

### 2.5 Statistical Analysis

Demographic characteristics were compared between groups by Fisher’s Exact Test for Count Data and Kruskal-Wallis rank sum test, as Shapiro-Wilk test for normality showed nonparametric distribution of the data. Tests were performed with an alpha of 0.05 to interrogate the relationship between CpG methylation and SNP genotype. Multiple comparisons were measured with the Bonferroni method to further elucidate differences between groups. Analyses were conducted in R v4.4^37^.

## 3. Results

### 3.1 Demographic and Genetic Covariate Comparisons

Clinical and demographic characteristics did not significantly differ between the three groups besides more never smokers (77% vs. 33-48%) and lower BMI (26.9 vs. 29.0-31.5 kg/m^2^) in CON (**Table 1**). Genetic measurements such as principal components (PCs) and polygenic risk scores (PRSs) were also compared between groups. None of the PCs or PRSs were statistically different between groups apart from the expected difference in BD PRS between BD groups and CON.

**Table 1.**
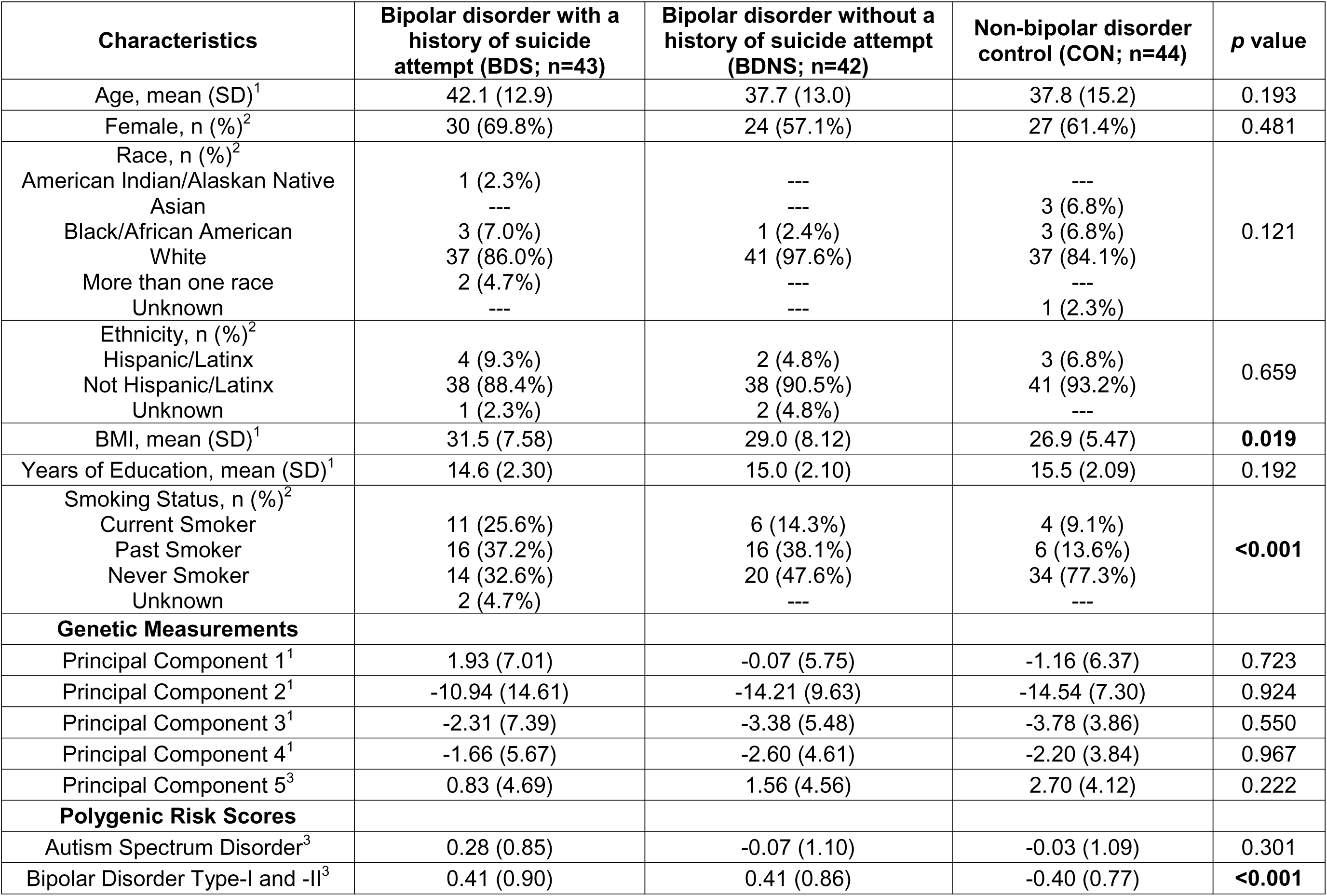
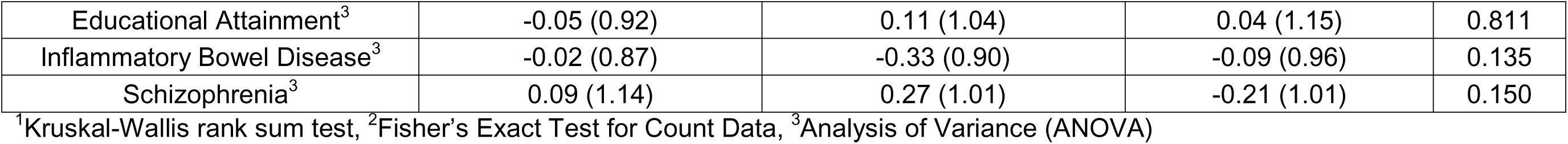
Demographic Characteristics of Groups.

### 3.2 DNAm Analysis

*ARHGEF38* was not significantly differentially methylated in any group at any CpG site investigated. However, for each CpG site, distinct DNAm clusters were noted (**Figure 2**). Covariate analyses were conducted to ascertain if these clusters were driven by demographic features previously listed in **Table 1**, however, none proved to drive this distinction (data not shown).

**Figure 2.**
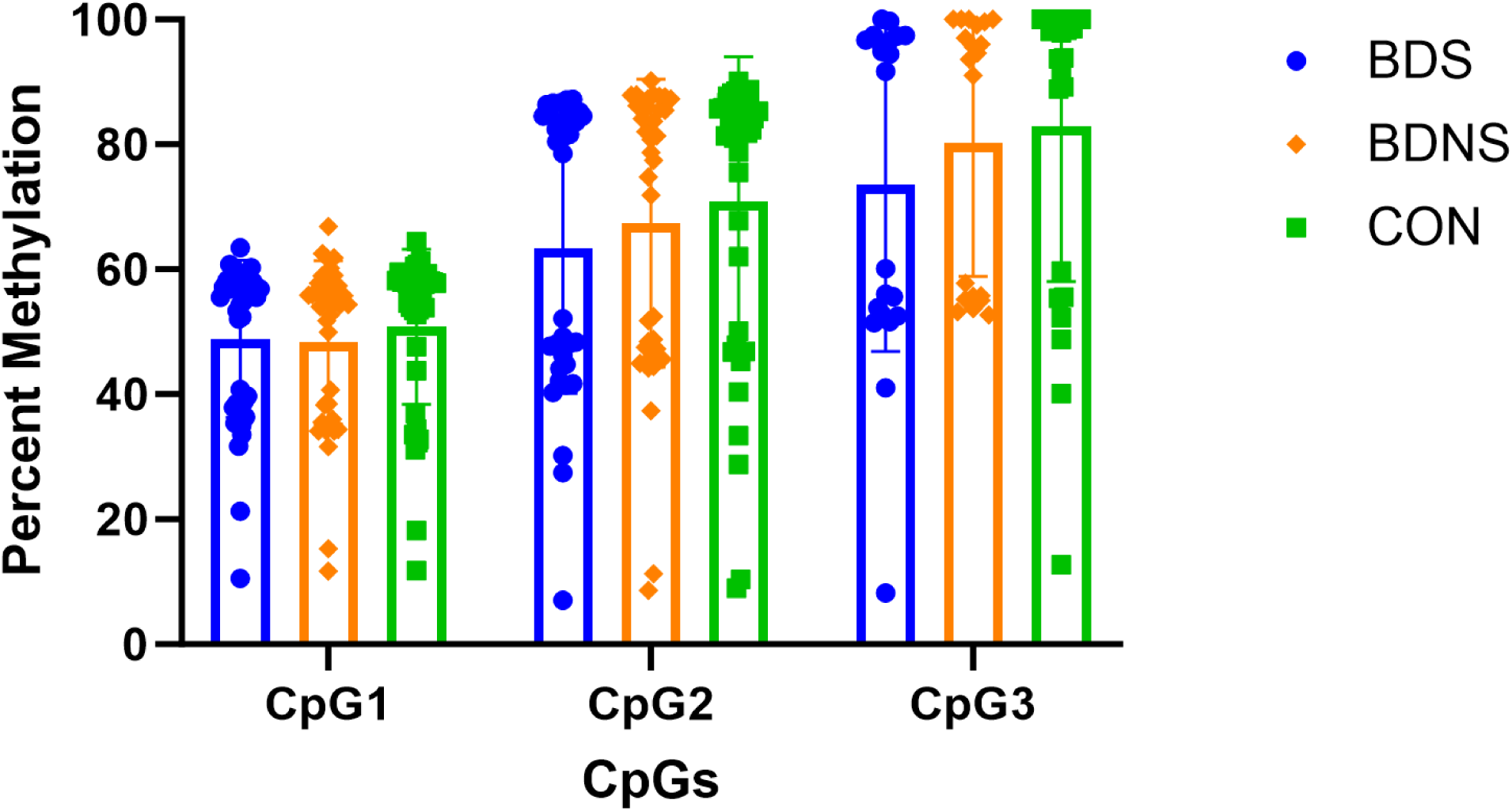
*ARHGEF38* methylation at each CpG site by group. CpG1 (chr4:106553213-106553214), CpG2 (chr4:106553273-106553274), CpG3 (chr4:106553280-106553281). All genomic locations use hg19 genome assembly. BDS; bipolar disorder with history of suicide attempt, BDNS; bipolar disorder without history of suicide attempt, and CON; non-bipolar disorder controls.

### 3.3 Methylation vs Genotype Analysis

To investigate factors potentially influencing the methylation in these CpG sites, we integrated SNP data from the region of interest. The genotypes of rs2121558 and rs1447093 were associated with clusters of DNAm in CpG1, CpG2 and CpG3 (**Figure 3a, c-d, f-g**, and **i**). The reference genotype (e.g., A/A for rs2121558 and G/G for rs1447093) was associated with the highest methylation at each CpG site. Heterozygosity was associated with a lower DNAm than the reference genotype, and a homozygous alternate genotype was associated with the lowest DNAm level for each CpG site. These relationships were observed regardless of phenotype (**Figure 4**, **Table 2**). SMR found no effect of these SNPs on SA via brain DNAm (**Supplemental Table 1**).

**Figure 3.**
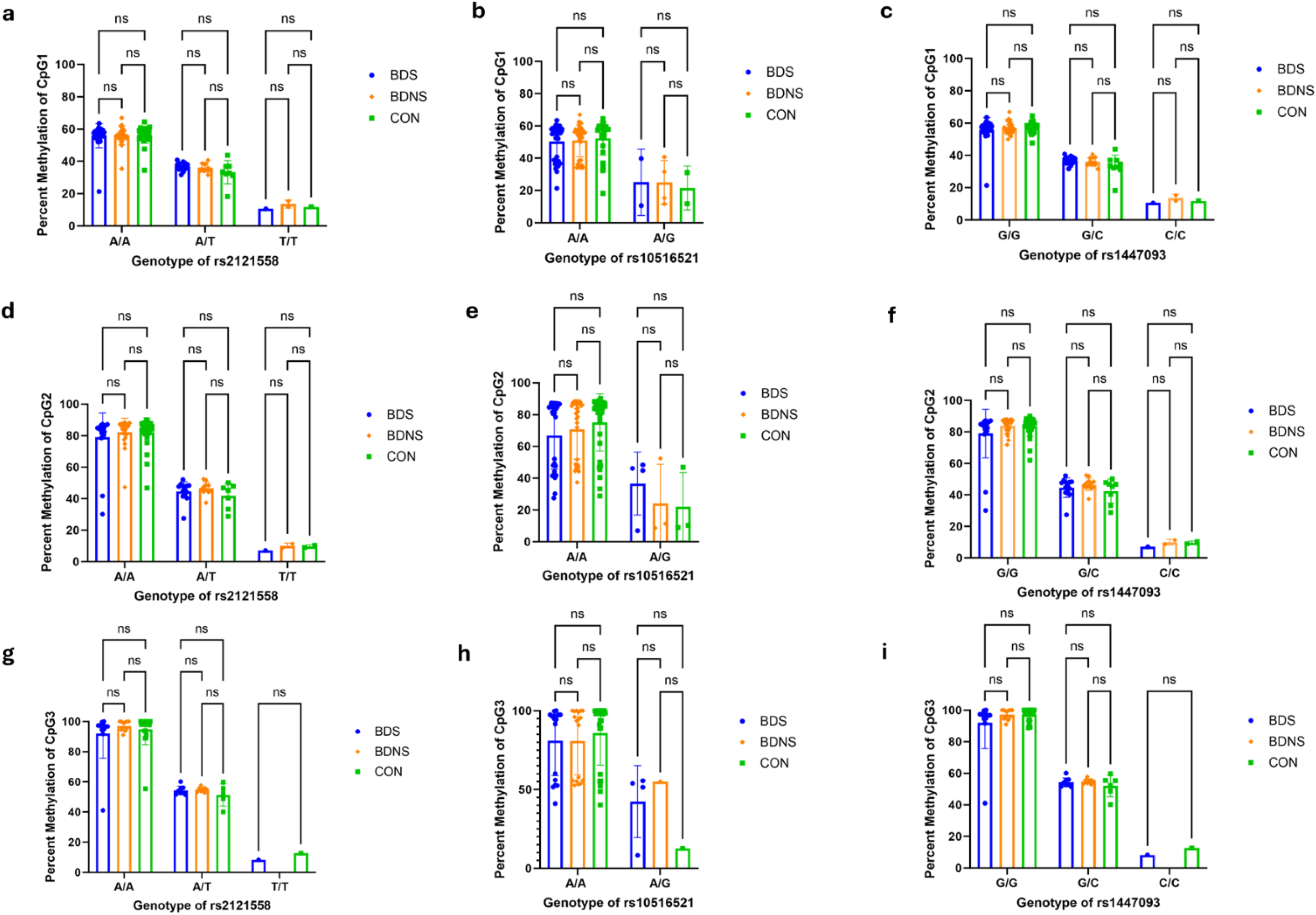
*ARHGEF38* methylation separated by genotype and group. Percentage methylation per genotype for CpG1 versus **a)** rs2121558, **b)** rs10516521, **c)** rs1447093; for CpG2 versus **d)** rs2121558, **e)** rs10516521, **f)** rs1447093; and for CpG3 versus **g)** rs2121558, **h)** rs10516521, and **i)** rs1447093. CpG1 (chr4:106553213-106553214), CpG2 (chr4:106553273-106553274), CpG3 (chr4:106553280-106553281). All genomic locations use hg19 genome assembly. BDS; bipolar disorder with history of suicide attempt, BDNS; bipolar disorder without history of suicide attempt, and CON; non-bipolar disorder controls. ns indicates not significant results.

**Figure 4.**
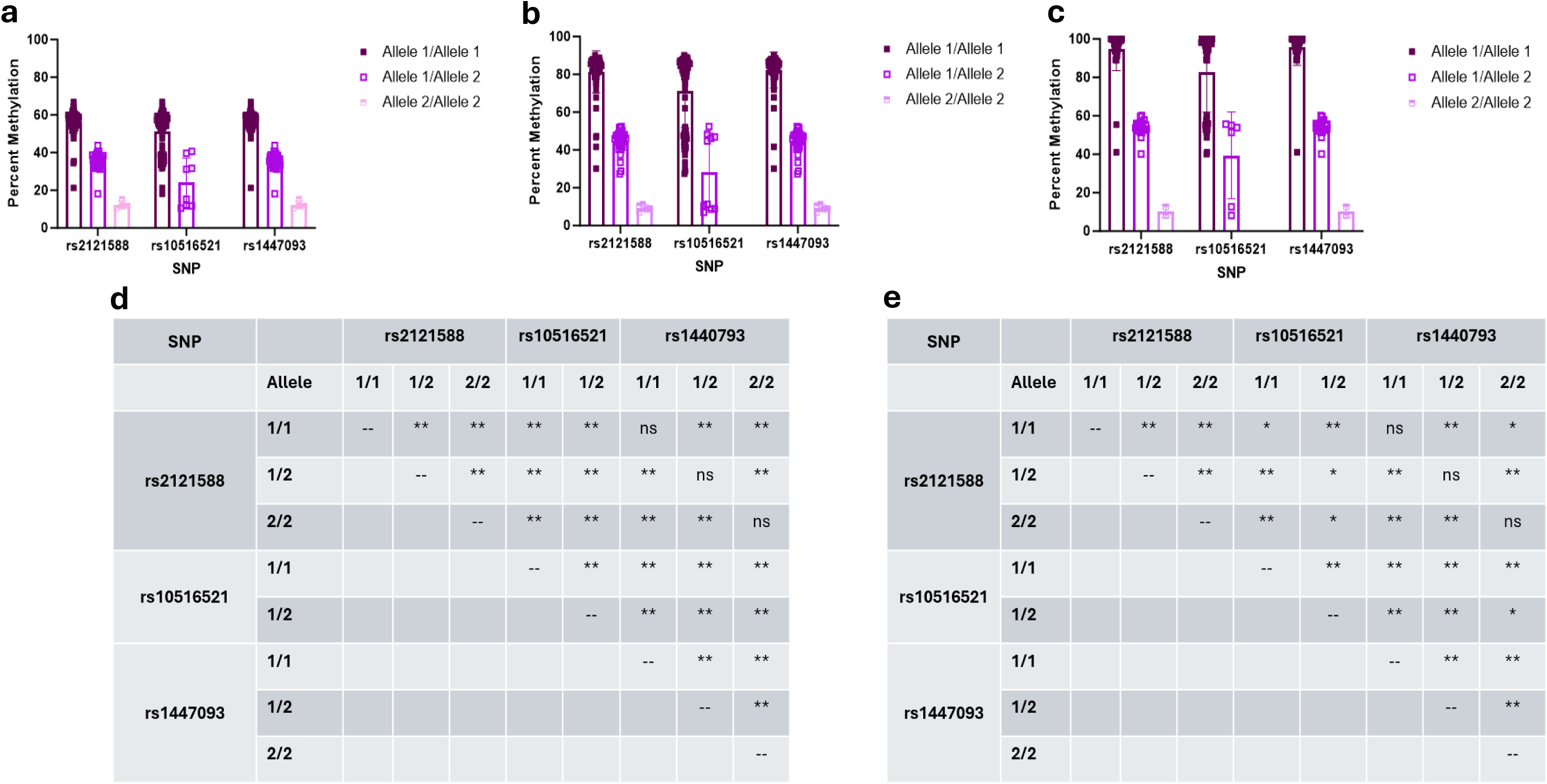
*ARHGEF38* methylation at each CpG site by genotype. Percentage Methylation for **a)** CpG1 versus genotype (Allele 1/Allele 1: reference genotype; Allele 1/Allele 2: heterozygous SNP, Allele 2/Allele 2: homozygous SNP), **b)** CpG2 versus genotype, and **c)** CpG3 versus genotype. Two-way ANOVA *p-*value results with Tukey’s multiple comparison test for **d)** CpG1 and CpG2 methylation, and **e)** CpG3 methylation comparing genotypes for each SNP. ** indicates *p* <0.001, ** indicates *p*<0.01, and ns indicates not significant results.

**Table 2.** Average Percentage Methylation per SNP-CpG Relationship.

| **BDS** | **rs2121558** |  |  | **rs10516521** |  |  | **rs1447093** |  |  |
| --- | --- | --- | --- | --- | --- | --- | --- | --- | --- |
|  | **A/A** | **A/T** | **T/T** | **A/A** | **A/G** | **G/G** | **G/G** | **G/C** | **C/C** |
| **CpG1** | 55.97<br>(7.39) | 36.58<br>(2.42) | 10.52<br>(--) | 49.97<br>(11.07) | 30.45<br>(13.45) | N/A | 57.31<br>(2.61) | 35.62<br>(4.48) | 10.52<br>(--) |
| **CpG2** | 80.37<br>(13.06) | 47.34<br>(11.67) | 7.02<br>(NA) | 70.27<br>(21.33) | 36.60<br>(19.78) | N/A | 81.85<br>(15.48) | 46.99<br>(6.16) | 7.02<br>(0.0) |
| **CpG3** | 93.02<br>(13.05) | 57.06<br>(11.35) | 8.17<br>(--) | 82.21<br>(20.79) | 42.29<br>(22.81) | N/A | 94.42<br>(11.02) | 57.01<br>(10.96) | 8.17<br>(--) |
| **BDNS** | **rs2121558** |  |  | **rs10516521** |  |  | **rs1447093** |  |  |
|  | **A/A** | **A/T** | **T/T** | **A/A** | **A/G** | **G/G** | **G/G** | **G/C** | **C/C** |
| **CpG1** | 56.20<br>(5.35) | 36.09<br>(2.60) | 13.52<br>(2.54) | 50.90<br>(10.13) | 24.86<br>(13.69) | N/A | 56.96<br>(3.55) | 36.04<br>(2.50) | 13.52<br>(2.54) |
| **CpG2** | 82.41<br>(8.00) | 46.48<br>(3.93) | 9.95<br>(1.90) | 72.80<br>(17.76) | 29.63<br>(22.89) | N/A | 83.71<br>(4.14) | 46.54<br>(3.77) | 9.95<br>(1.90) |
| **CpG3** | 95.40<br>(7.75) | 54.56<br>(1.43) | 14.61<br>(0.78) | 84.76<br>(19.38) | 34.45<br>(22.93) | N/A | 96.81<br>(2.15) | 54.78<br>(1.57) | 14.61<br>(0.78) |
| **CON** | **rs2121558** |  |  | **rs10516521** |  |  | **rs1447093** |  |  |
|  | **A/A** | **A/T** | **T/T** | **A/A** | **A/G** | **G/G** | **G/G** | **G/C** | **C/C** |
| **CpG1** | 56.44<br>(4.99) | 33.19<br>(7.26) | 12.51<br>(1.01) | 52.28<br>(10.30) | 18.71<br>(10.76) | N/A | 57.11<br>(3.18) | 33.33<br>(6.80) | 12.51<br>(1.01) |
| **CpG2** | 82.38<br>(8.40) | 42.22<br>(7.51) | 9.65<br>(1.03) | 75.41<br>(17.59) | 22.05<br>(21.49) | N/A | 83.46<br>(5.66) | 42.73<br>(7.19) | 9.65<br>(1.03) |
| **CpG3** | 95.74<br>(7.67) | 52.28<br>(5.79) | 13.39<br>(1.01) | 88.29<br>(18.19) | 26.77<br>(23.19) | N/A | 96.96<br>(2.87) | 52.62<br>(5.51) | 13.39<br>(1.01) |

Mean percentage methylation at each CpG site per SNP genotype. Standard deviation presented in paratheses. N/A indicates that the genotype does not exist in the group. BDS; bipolar disorder with history of suicide attempt, BDNS; bipolar disorder without history of suicide attempt, and CON; non-bipolar disorder controls.

The SNPs rs2121558 and rs1447093 influence *ARHGEF38* DNAm in a genotype-dependent manner as a pattern of high, medium, and low DNAm correlates with distinct genotypes. However, this pattern is not seen with rs10516521 (**Figure 3b, e,** and **h**). Instead, DNAm clusters are seen within genotype groups (for example ∼100% and ∼50% DNAm subgroups with the A/A genotype; **Figure 3h**). Covariate analysis did not uncover a demographic covariate driving this observation (data not shown). This would indicate that while rs2121558 and rs1447093 may be influencing the DNAm seen at each of the three CpG sites, rs10516521 is not associated with *ARHGEF38* DNAm.

meQTL analyses were performed looking at all common SNPs on chromosome 4 with the three SNPs, demographic variables, and genetic variables used as covariates to interrogate potential causal SNPs for the pattern of DNAm. SNPs on *GABRB1, ATP10D,* and PARM1 may influence CpG1, SNPs on *SLC7A11, LINC00499, GRID2, KCNIP, PRDM5,* and LNX1 may influence CpG2 DNAm, and SNPs on *ANXA10, F11-AS1, LINC02232, NR3C2,* and *SPOCK3* may influence CpG3 DNAm. We only report SNP-CpG pairs with Bonferroni corrected *p*-values <5.0E-04 to reduce false positives (**Figure 5**, **Table 3**, and **Supplemental Table 2**).

**Figure 5.**
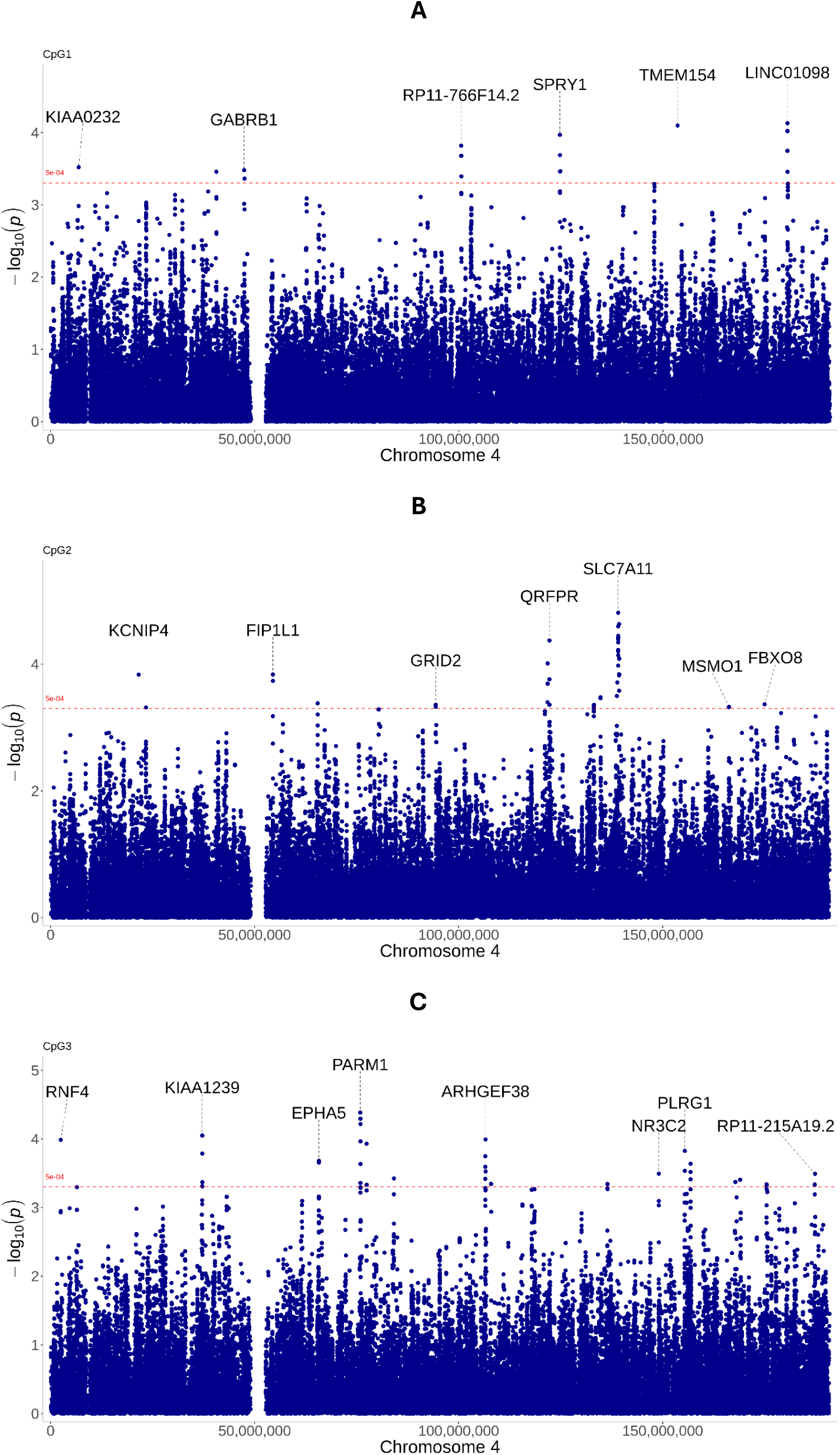
Manhattan plots of the meQTL associations for each CpG site. Manhattan plots of the meQTL associations in whole-blood samples for **a)** CpG1, **b)** CpG2, and **c)** CpG3. The line represents the significance level at Bonferroni corrected p-value of 5.00E-04.

**Table 3.**
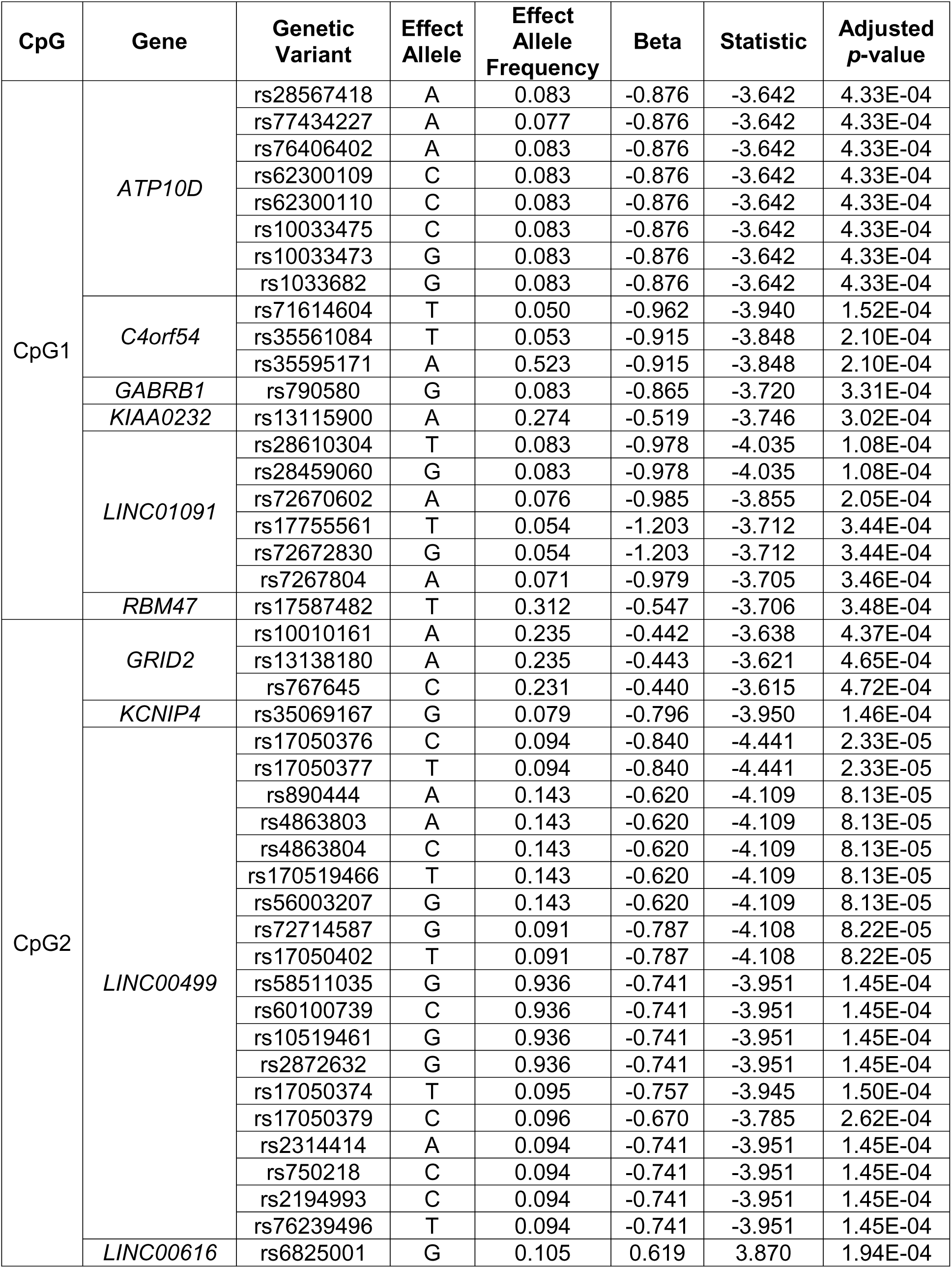
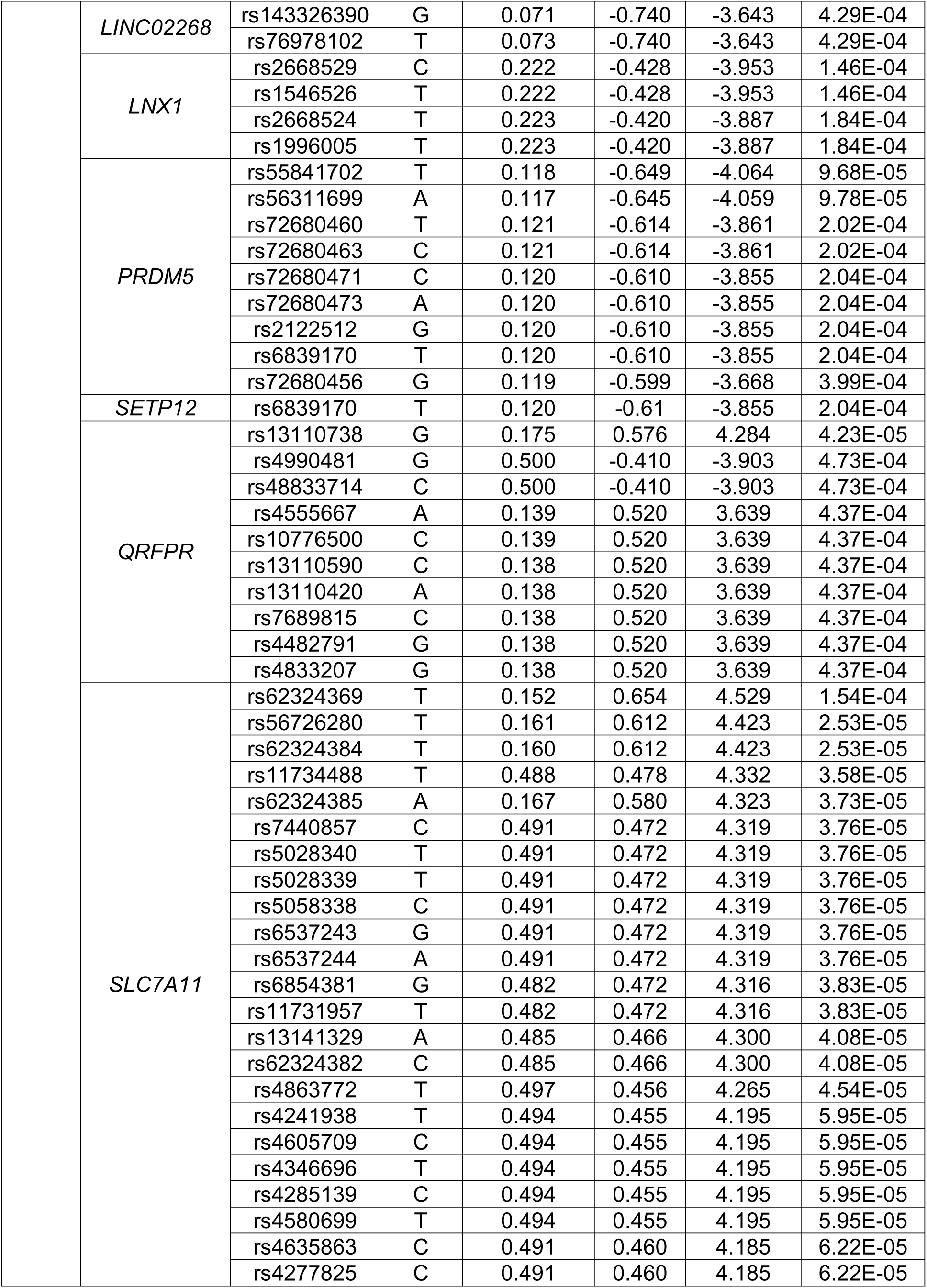
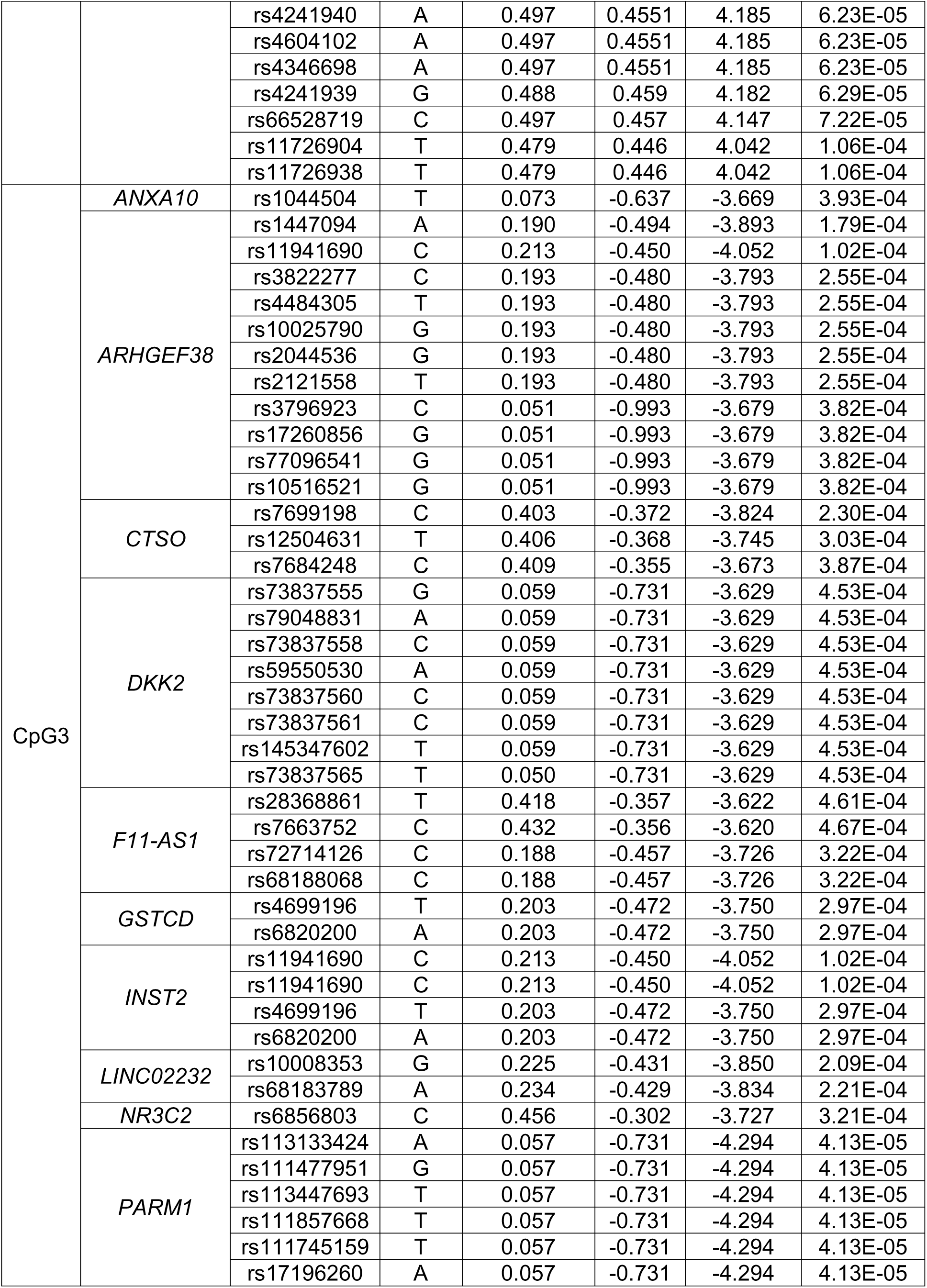
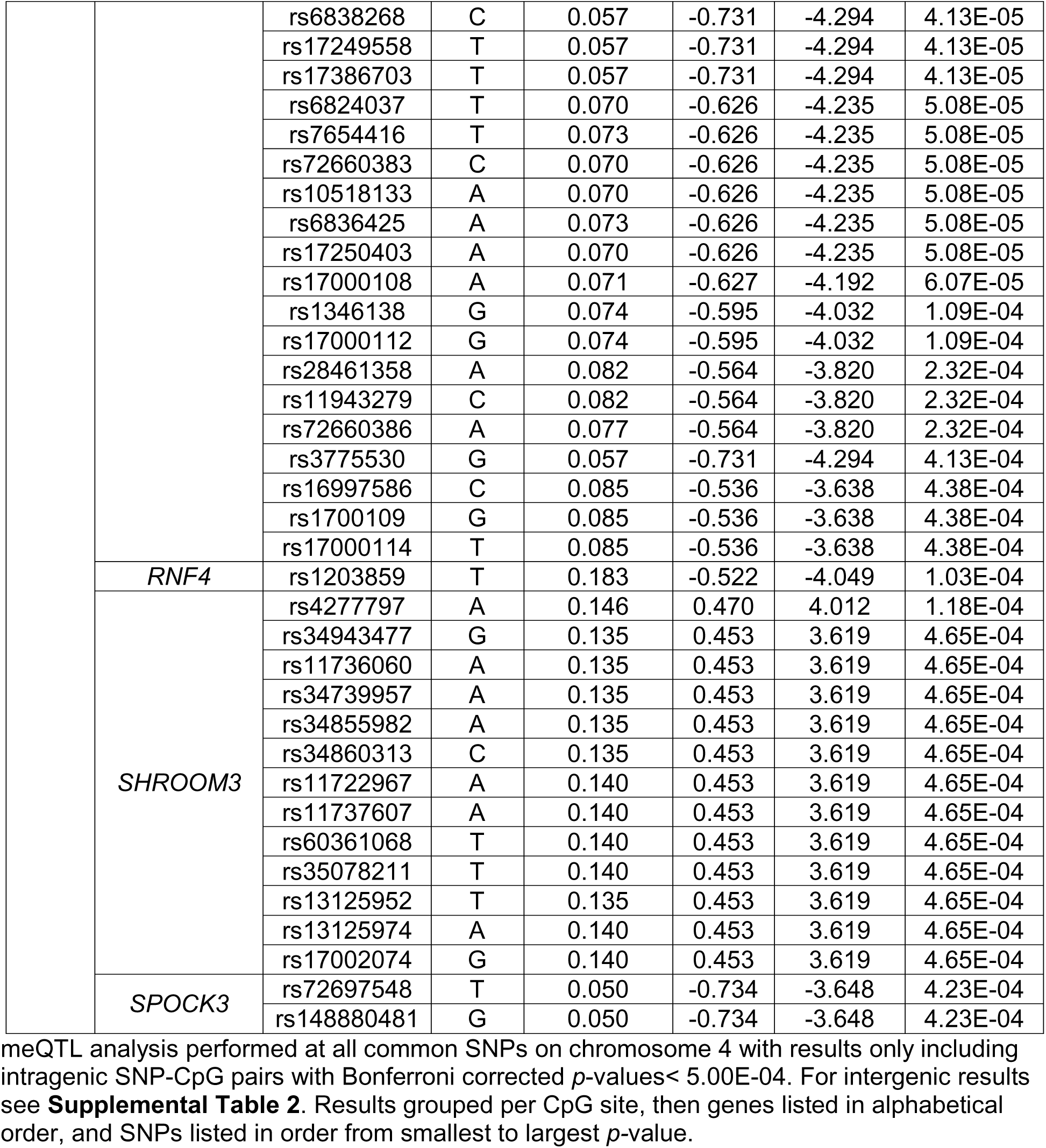
Intragenic meQTL analysis results.

## 4. Discussion

*ARHGEF38* is a relatively unknown gene associated with prostate^38,39^ and lung^40^ cancer, and non-syndromic cleft lip and/or cleft palate^41^, in addition to the SD study upon which this investigation is based^18^. *ARHGEF38* is not highly expressed in either the brain or whole blood^42^, suggesting that hypomethylation leading to increased expression may have functional relevance^18^. Replication studies are important to demonstrate generalizability and robustness, which has been a challenge for candidate gene approaches^43^. This SA study did not replicate the association between *ARHGEF38* and SD, however, there were limitations: the presence of genetic variation influencing DNAm, comparison of SA to SD, and use of whole blood versus brain tissue.

Our study highlights the importance of considering genetics when studying DNAm. We identified SNPs, rs2121558 and rs1447093, associated with specific DNAm clusters, suggesting regulation of DNAm in *ARHGEF38* by *cis-*SNPs. Notably, rs2121558 and rs1447093 are in higher linkage disequilibrium (LD; R^2^=0.982) than rs2121558 and rs10516521 (R^2^=0.164) or rs2121558 and rs1447093 (R^2^=0.163)^44^.

The ability to detect meQTLs depends on SNP allele frequency and LD, as well as CpG site variance in methylation, and therefore, the power of the SNP-CpG relationship^45^. Several studies have assessed functionally relevant meQTLs and psychiatric disorders^46–49^, however only one has looked at meQTLs in suicide attempt^50^. Our analyses focused on three targeted CpG sites in *ARHGEF38*. The methylation pattern of CpG1 could be informed by SNPs on *GABRB1*, a gene associated with schizophrenia^51^, major depressive disorder^52^, self-regulation disorders^53^, and risk-taking behaviors^54,55^, although we did not see these specific variants in our analysis. The SNPs predicted to influence CpG2 are in genes associated with depressive phenotypes, *SLC7A11*^56^ and *LINC00499*^57,58^; obsessive-compulsive disorder with *GRID2*^59^; suicidal thoughts and behaviors with *SLC7A11*^56^*, LINC00499*^60^, *KCNIP4*^61^, and *GRID2*^62^. Perhaps of most interest are the genes associated with DNAm, *PRDM5*^63^*, LNX1*^64^, and *KCNIP4*^61^. This suggests that DNAm of CpG2 may be informed by SNPs on genes already associated with regulation of DNAm. Other genes implicated in CpG3 methylation were associated with depressive symptomology, *ANXA10*^65^ and *F11-AS1*^66^, and schizophrenia, *LINC02232*^51,67,68^, *NR3C2*^69^ and *SPOCK3*^51^. To focus this study, we did not investigate intergenic SNPs that may also influence DNAm of the three targeted CpGs (**Supplemental Table 2**). Overall, this analysis highlights the complexity of SNP-CpG pairs, with SNPs on chromosome 4 implicated in epigenetic changes and even susceptibility for psychiatric disorders.

Our analyses also uncovered an association between CpG1 methylation in *ATP10D*, rs77434227-A, which has been implicated in ceramide metabolism^70^. Variants in *PARM1*, rs72660383-C and rs1051833-A, were associated with methylation of CpG3, which have been associated with calcium measurements^71^ and childhood steroid-sensitive nephrotic syndrome^72^ respectively. As *ARHGEF38* is predicted to be involved in intracellular signal transduction^41^, these variants could modify *ARHGEF38*’s ability to bind with lipids and calcium to alter membrane targeting and signaling.

Another key difference between the prior study and this one was the phenotype of SB measured. In this study, we focused on SA whereas the original study focused on suicide decedents. Historically, research focused on SB, which encompassed SA and SD. Although SA and SD share clinical associations, recent studies show biological distinctions between the two^73,74^, suggesting caution when comparing them in epigenetic studies.

Finally, it is important to consider the cell populations being investigated. Surrogate tissue, such as whole blood, has been used to explore DNAm in psychiatric diseases due to the inaccessible nature of the brain^25^. Brain and blood DNAm patterns are highly correlated, with one study showing a correlation of *r* =0.86 across subjects, while within subjects, blood had the highest proportion of nominally significant CpGs correlated to the brain when compared with other surrogate tissues, such as buccal tissue or saliva^75^. Unfortunately, we could not determine the correlations within our chosen *ARHGEF38* CpG sites as they were not covered by the Illumina EPIC array from the comparison study^75^, which found several cell-type specific CpG sites. This suggests that the blood methylome may not be a consistent proxy for the brain methylome.

In conclusion, our study found that *ARHGEF38* DNAm is not associated with SA in individuals with BD. Not only does this show the inherent differences of methylation between SD vs SA, but the CpG-SNP relationships uncovered in this study emphasize the important relationship between genetics and epigenetics when studying DNAm.

## Data Availability

All data produced in the present study are available upon reasonable request to the authors

## Acknowledgments

We thank the study participants for their willingness to participate in the study and Hsiang Wen for sample collection and management. We also acknowledge the Iowa NeuroBank Core, particularly Queena Lin and Mara Jendro, for access to the PyroMark Q48.

